# Physiologically relevant real-world light exposure and its behavioural and environmental determinants in Kumasi, Ghana

**DOI:** 10.1101/2025.09.11.25334564

**Authors:** Gabriel Kwaku Agbeshie, Isaiah Osei Duah, Albert Kwadjo Amoah Andoh, Josephine Ampong, Nana Akwasi Owusu Mensah, Awurama Yenkua Ampoma-Mensah, Johannes Zauner, Manuel Spitschan, Kwadwo Owusu Akuffo

## Abstract

Light plays a significant role in human physiology and behaviour, influencing sleep, mood, alertness and overall health. Yet light exposure remains a neglected determinant of health, with most evidence coming from high-income countries. This data note presents a dataset collected in Kumasi, Ghana, between October 2024 and February 2025, containing personal light exposure measured with wearable melanopic light loggers alongside behavioural and environmental factors obtained through self-reports. Fifteen participants (n=8 female, mean±SD age 22.6±1.2 years) wore three ActLumus light-logging devices continuously for seven days, recording light exposure every 10 seconds at the near-corneal plane, chest and wrist. Participants also completed daily questionnaires on sleep, mood and physical activity, as well as structured assessments of their sleep environment and light-related behaviours. This dataset provides the first high-resolution account of daily light exposure in sub-Saharan Africa. By enabling cross-context comparisons, it contributes to understanding the diversity of light environments globally and can inform interventions for sleep health, mental health and chronic disease prevention.

**Plain language summary:** Light affects how we sleep, feel and stay healthy. But most research on light exposure has been carried out in high-income countries, meaning we know very little about people’s real-life light environments in other parts of the world. This study provides the first detailed dataset of daily light exposure in sub-Saharan Africa. Fifteen young adults in Kumasi, Ghana, wore small devices that measured the light around them every few seconds for one week, while also answering short questions about their sleep, mood and daily routines. The dataset shows how people in Ghana experience light in everyday life and allows comparisons with data from other countries. By making this resource openly available, we hope to support future research on how light exposure influences sleep, mental health and the risk of long-term diseases — helping to shape healthier light environments worldwide.

## Introduction

Light exposure is an important environmental factor that significantly influences human health and well-being [1]. In addition to its central role in vision, light acts as the primary zeitgeber, synchronising the endogenous circadian rhythms with the natural light-dark cycle [2]. However, the prevalence of artificial lighting and modern indoor lifestyles can disrupt this synchronization, leading to various health issues due to altered daily patterns of light exposure [3–6]. The non-visual effects of light involve specialized photoreceptors in the eye and are mainly mediated by the intrinsically photosensitive retinal ganglion cells (ipRGCs), which are most sensitive to short-wavelength light [7, 8].

Analysing real-world light exposure and related behaviours is key for a better understanding of and potential reduction in any negative health consequences arising from light at the ‘wrong’ time, while enhancing the acute and short-term benefits of light exposure, such as boosts in alertness and improved sleep quality. Although extensive laboratory research has characterised the effects of light under controlled conditions, these light stimuli rarely reflect the light exposures encountered under real-world conditions [9, 10]. Empirical studies of light exposure in daily life using wearable light loggers are essential for providing additional information about exposure patterns and their potential consequences for health.

Wearable light loggers and dosimeters have emerged as valuable tools for objectively measuring personal light exposure in real-world settings [11–14]. These devices can capture spectral information across the visible range and output various light exposure metrics, including visual quantities (e.g. photopic illuminance) and non-visual quantities (e.g. alpha-opic irradiance, melanopic equivalent daylight illuminance, abbreviation as melanopic EDI) [15, 16], which in turn can be aggregated into relevant summary metrics [17–19]. Objective measurements can be supplemented by subjective tools that capture light-exposure-related behaviours, providing a more comprehensive understanding of how individuals interact with their light environment [20].

Despite a growing interest in personal light exposure, more extensive data is needed from diverse populations and geographical locations to fully understand variability in light exposure patterns and their health implications. To date, most research has focused on populations in Europe or North America [17, 21]. First, comparisons with other regions of the world demonstrate the differences between countries, culturesand climate conditions, as a recent study on light exposure in Malaysia and Switzerland has shown [22]. Datasets from other regions are less common and even fewer are publicly available, which limits our ability to assess generalisability and identify population-specific factors that influence light exposure and its effects on human health. Light exposure data collected in Ghana provides an opportunity to study how different environmental conditions, cultural practices and daily routines correlate with the light exposure patterns. This contributes to a broader understanding of the diversity of human physiological responses to light, particularly within the African population.

This data note describes a dataset on personal light exposure collected in Ghana. The primary objective of the dataset was to characterize personal light exposure patterns in Ghana and investigate the real-world determinants of participant’s exposure. The use of a standardized, multi-site study protocol [23] ensures that the data are comparable with a growing number of other sites around the world including Germany, the Netherlands, Sweden, Spain, Turkey and Costa Rica. By making this dataset publicly available and adhering to the principles of FAIR (findable, accessible, interoperable, reusable) data [24], we aim to contribute valuable information to the field, facilitating further research and the development of more personalised and effective interventions for optimising light exposure for health and well-being.

## Methods

This dataset was collected in an observational field study in which participants were recruited in Kumasi, Ashanti Region, Ghana (coordinates 6.6750074282377385 N, -1.572643823555129 W), to evaluate their personal light exposure patterns using wearable light loggers and questionnaires following a standardized, multi-site protocol [23]. This study was conducted within the framework of the Metrology for wearable light loggers and optical radiation dosimeters (MeLiDos) project that aims to quantify individual light exposure using wearable light loggers and solar UV dosimeters [25]. For the general overview of this study, **see Figure 1**.

**Figure 1:**
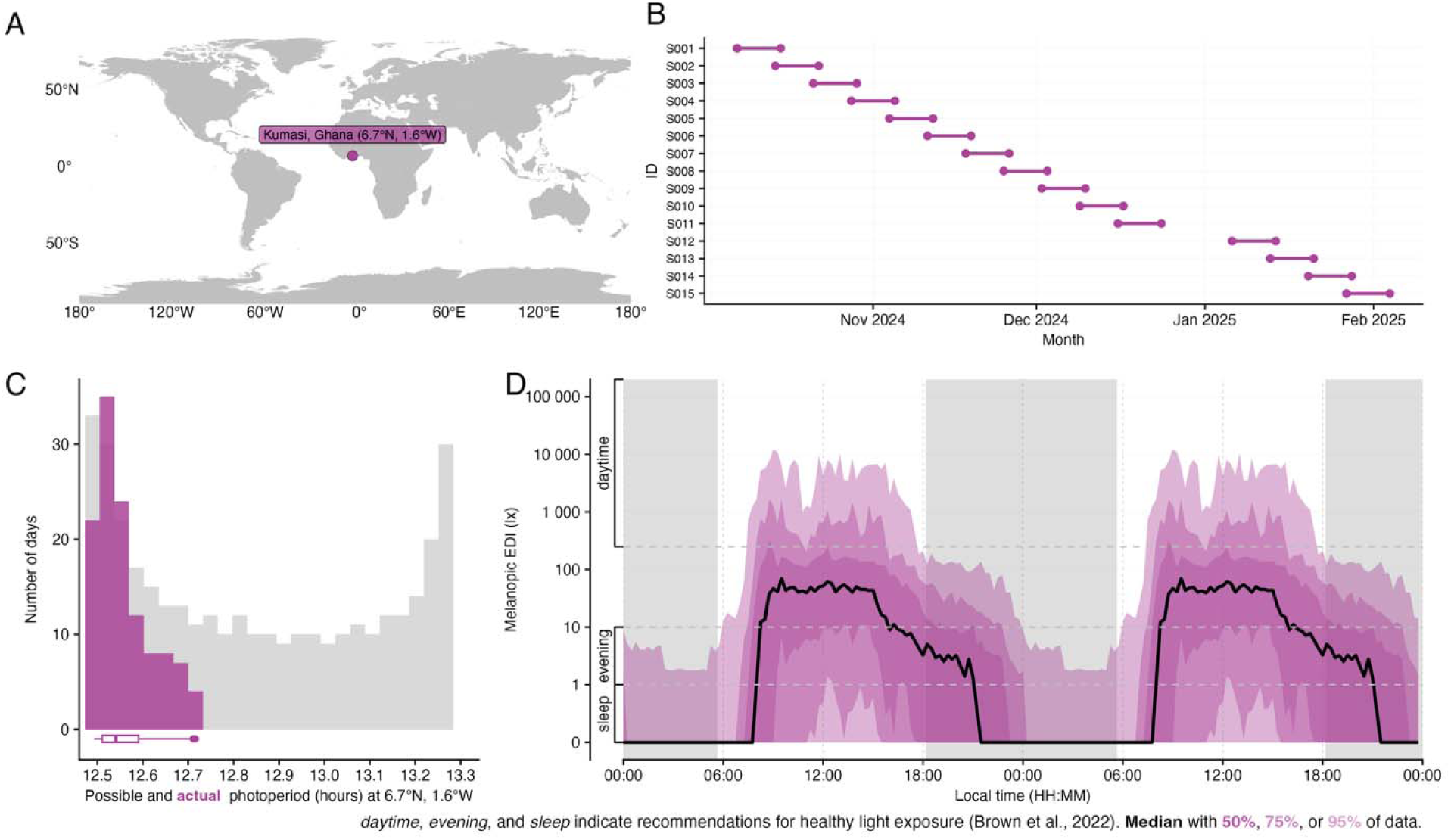
An overview of the study design. **A**, Study location shown on map. **B**, Participants recording periods including implicit missing data in grey. Of note, the gap in between participant S011 and S012 is because of a week break during the data collection during the winter holiday period. **C**, Actual photoperiod duration in Kumasi, Ghana (pink). **D**, Recommended healthy daytime, evening and sleep light exposure (melanopic equivalent daylight illuminance, melanopic EDI).

### Participant recruitment

Participants were recruited via advertisements disseminated as flyers on various social media platforms. Participants were directed to scan a QR code to access an online platform (Research Electronic Data Capture, or REDCap [26, 27]) on which they could complete the first screening survey. The aims of the study and detailed information were provided during this screening step. Eligible participants were aged 18–65 years, had no psychiatric or sleep disorder, were not using tobacco or recreational drugs or taking medication, had normal vision and were not using prescribed glasses and lived within a 60-km radius of the research centre. Eligible participants were sent a picture of a spectacle-mounted ActLumus light logger (Condor Instruments, São Paulo, Brazil) and were asked to confirm that they would feel comfortable wearing it throughout the study period. Participants received financial remuneration of €82.30 at the end of the study based on their compliance with the experimental light study, which required them to wear the light logger for at least 80% of their waking hours. 15 participants were recruited for the study.

### Procedures

Eligible participants were invited to the department on a given Monday to begin the light exposure assessment and finished the following Monday. On the first Monday (day 1), participants received detailed information about the study and signed an informed consent form. They also received three wearable light loggers, which they were instructed to wear at chest level, at the near-corneal plane and on their wrist. They were given instructions on how to use the devices correctly. They also installed the MyCap app [28], which integrates with REDCap and was used to complete the daily questionnaires during the light study.

### Measures and data collection

#### ActLumus light loggers (Condor Instruments, São Paulo, Brazil)

Three ActLumus light loggers (Condor Instruments, São Paulo, Brazil) were used to objectively measure personal light exposure and activity at different locations. One ActLumus was positioned near the corneal plane on a 3D-printed holder attached to the bridge of a pair of non-prescription glasses. A second light logger was worn as a manufacturer-supplied pendant attached to a lanyard to measure light at chest level. The third light logger was worn at wrist level using the manufacturer-provided wristbands. The devices record across the visible spectrum and provide calibrated alpha-opic and photopic metrics, as well as movement using an integrated tri-axial accelerometer. Each light logger was set to a 10-second sampling interval to achieve highly temporally resolved data. Participants were also instructed to follow instructions about when to wear and remove them and how to document wear and non-wear. They were also informed to ensure that the light loggers were not obstructed by clothing and to remove them when in contact with water or during intense sporting activities. The devices were never charged or turned off during the light study week. The light exposure data were retrieved from the devices when they were returned on the final Monday (day 8) and stored on a secure, personal computer on campus.

### Chronotype questionnaires

On the first day of the light exposure study, participants completed the chronotype questionnaire. This consisted of the Munich Chronotype Questionnaire (MCTQ) [29], which assesses circadian time using questions about sleep and wake habits during work and leisure periods and the Morning-Eveningness Questionnaire (MEQ) [30] to determine their circadian preference, i.e. the times of day at which they perform certain activities.

### Visual Light Sensitivity Questionnaire-8 (VLSQ-8)

Participants completed the eight-point Visual Light Sensitivity Questionnaire (VLSQ-8) [31] on the final day of the data collection week after returning the wearable devices. The questionnaire asked participants to estimate the frequency and severity of their photosensitivity, as well as its impact on their daily behaviours, using a five-point Likert scale (1 = “Never” to 5 = “Always”).

### Morning sleep log

Participants completed the Sleep Diary every morning after waking up in order to assess their sleep timing, duration and quality. The questionnaire consisted of nine items, with the final item scored on a five-point Likert scale ranging from 1 (“Very poor”) to 5 (“Very good”).

### Ecological momentary assessment (“Current conditions”)

Participants completed a short questionnaire four times a day (at 11:00, 14:00, 17:00 and 20:00) about their current light environment, mood and level of sleepiness. They received reminders via the REDCap/MyCap app and were instructed to set their own phone alarms to help them remember. For the light conditions section, participants selected one or two main light sources from a list of eight options based on a modified version of the Harvard Light Exposure Assessment Diary (H-LEA). Mood was measured using a shortened version of the Mood Zoom questionnaire [32] and sleepiness was rated using a 10-point Karolinska Sleepiness Scale (KSS) [33], which ranges from 1 (“Extremely alert”) to 10 (“Extremely sleepy, fighting sleep”).

### Exercise log

Each evening before going to bed, the participants completed a custom-made questionnaire about their daily physical activity. The questionnaire collected information on the intensity of the exercise (vigorous, moderate or light), where it took place (indoors or outdoors) and how much time was spent sitting or lying down during the day.

### Wellbeing log

Each evening before going to sleep, the participants completed a modified version of the WHO-5 Wellbeing Index [34]. This comprised five statements about their mood, energy levels, sleep quality and interest in daily life. For four of the statements, participants rated how often they experienced positive emotions using a five-point Likert scale ranging from 0 ("Never") to 5 ("Always"). For the question about sleep quality, they rated it from 1 (“Very poor”) to 5 (“Very good”).

### Light exposure and activity log

Each evening, the participants completed the modified Harvard Light Exposure Assessment (H-LEA [35]), which was provided during their first visit to the office (on day 1). For each hour of the day, they recorded the main light source to which they were exposed, defined as the ’biggest and brightest light source’, as well as the activity in which they were engaged during that hour. Light sources were chosen from eight predefined categories, including indoor and outdoor electric light, daylight indoors or outdoors, screen-based light, darkness and light during sleep. If they were exposed to multiple light sources within the same hour, participants selected from a list of combined options. Activities were selected from eight categories, such as sleeping, working, commuting or spending time outdoors. To ensure compliance, participants submitted a photo of the completed form each morning to a personal shared folder on Google Drive. They also used the MyCap app to rate their confidence in the accuracy of their responses on a five-point Likert scale ranging from 1 (“Not confident at all”) to 5 (“Completely confident”).

### Light Exposure Behaviour Assessment (LEBA)

At the end of the study (on day 8), the participants completed the Light Exposure Behaviour Assessment (LEBA [20]), which is a 22-item questionnaire designed to assess light-related behaviours retrospectively during the light study week. The first three items, relating to the use of blue-filtering, orange-tinted, or red-tinted glasses, were excluded as these were not relevant due to the participants wearing light loggers. The final version included 19 items focusing on behaviours such as daylight exposure, smartphone use, bedtime light habits and electric light use at home. Participants rated how often they engaged in these behaviours using a five-point Likert scale ranging from 1 (“Never”) to 5 (“Always”).

### Assessment of Sleep Environment (ASE) questionnaire

On the final day (day 8), participants completed the 13-item Assessment of Sleep Environment (ASE [36]) questionnaire, which asked about factors in their sleeping environment, such as light, noise, temperature and humidity, that could influence sleep quality or affect light measurements from the light logger placed near them during sleep (e.g. light entering through windows). They rated their agreement with each statement on a five-point Likert scale ranging from 1 (strongly agree) to 5 (strongly disagree).

### Data processing and availability

The data collected in REDCap was exported as a comma-separated (CSV) file and processed using Microsoft Excel 2016. To anonymise the data, each participant was given a unique identifier ranging from KNUST_S001 to KNUST_S015and the position of the light logger was labelled as “w” for the wrist, “c” for the chestand “h” for the head. This dataset is publicly available on Zenodo under https://doi.org/10.5281/zenodo.15576732 [37]. While the repository contains the raw (anonymized) data at start, it will be augmented into a standard format at a later point. This augmentation includes the addition of metadata in a human- and machine-readable format as well as the correction and documentation of implausible entries by participants (e.g., mixing up AM and PM when entering times so that typical sequences like going to bed, sleeping, waking up, getting up, do not fall on a continuous timeline). All steps will be archived on Zenodo and assigned a persistent identifier (DOI) for full traceability. All data are available under the terms of the <u>Creative Commons Zero v1.0 Universal</u> (CC0 1.0).

### Dataset description

The dataset is organised into two main categories: Group (containing files for all participants) and Individual (containing participant-specific files and logs). Each participant is identified by a unique participant ID (PID). The study was carried out over seven days for each participant (eight calendar dates), with data collected using continuous wearable light loggers (ActLumus) and standardised questionnaires in RedCap via the MyCap mobile app. Summary data are presented in **Table 1**.

**Table 1.** Summary statistics of this dataset.

| Summary table |  |
| --- | --- |
| Kumasi, Ghana (6.7°N, 1.6°E), TZ: Africa/Accra |  |
| **Overview** |  |
| Participants | **15** |
| Participant-days | **120** days |
| ≥80% coverage | **88** days |
| Missing/irregular | **16.88%** (12.24% - 37.92%) |
| Photoperiod | **12h 33m** (12h 29m - 12h 42m) |
| **Metrics** |  |
| Dose | **10,307** ±12,469 (28 - 60,588) lx·h |
| Duration above 250 lx | **1h 43m** ±1h 49m (0s - 7h 45m) |
| Duration within 1-10 lx | **3h 22m** ±2h 33m (14m - 13h 19m) |
| Duration below 1 lx | **12h 48m** ±3h 35m (4h 8m - 22h 43m) |
| Period above 250 lx | **28m 12s** ±38m 22s (0s - 4h 6m) |
| Duration above 1000 lx | **56m 58s** ±1h 4m (0s - 5h 11m) |
| First timing above 250 lx | **08:59** ±02:00 (01:38 - 15:05) HH:MM |
| Mean timing above 250 lx | **12:07** ±01:36 (08:30 - 15:55) HH:MM |
| Last timing above 250 lx | **16:30** ±02:55 (08:53 - 23:16) HH:MM |
| Brightest 10h mean<sup>†</sup> | **71.1** ±118.8 (0.1 - 838.7) lx |
| Brightest 10h midpoint | **13:05** ±01:58 (04:59 - 18:59) HH:MM |
| Darkest 5h mean<sup>†</sup> | **0.0** ±0.0 (0.0 - 0.2) lx |
| Darkest 5h midpoint | **03:38** ±04:15 (02:29 - 23:35) HH:MM |
| Interdaily stability | **0.276** ±0.062 (0.162 - 0.364) |
| Intradaily variability | **1.390** ±0.399 (0.636 - 1.923) |
| values show: **mean** ±sd (min - max) and are all based on measurements of melanopic EDI (lx) |  |
| <sup>†</sup> Values were log 10 transformed prior to averaging, with an offset of 0.1, and backtransformed afterwards |  |

### Group-level folder

This folder contains structured data to facilitate import during data analysis (see **Table 2**).

**Table 2.** Description of group-level folders.

| Folder Name | Description |
| --- | --- |
| **chronotype** | Four CSV files: one for the Morningness-Eveningness Questionnaire (MEQ), another for the Munich Chronotype Questionnaire (MCTQ), lookup tables for MEQ and MCTQ, each containing N = 15 responses |
| **demographics** | Two CSV files: one with demographics data only and another with lookup table for all participants. |
| **discharge** | Two CSV files: one containing responses to five post-study questionnaires: LEBA, ASE, VLSQ-8, mTFA and general feedback; the other with lookup table for 15 participants |
| **screening** | Two CSV files: one with both demographics and health screening data for all participants and demographics and health screening lookup table. |

LEBA, Light Exposure Behaviour Assessment; ASE, Assessment of Sleep Environment; VLSQ-
8, Visual Light Sensitivity Questionnaire-8; mTFA, Modified Theory Framework of Acceptability

### Individual-level folder

This contains the 15 participants’ folders with subfolders named by domain. The individual files reflect the data collection schedule and instruments used (see **Table 3**).

**Table 3.**
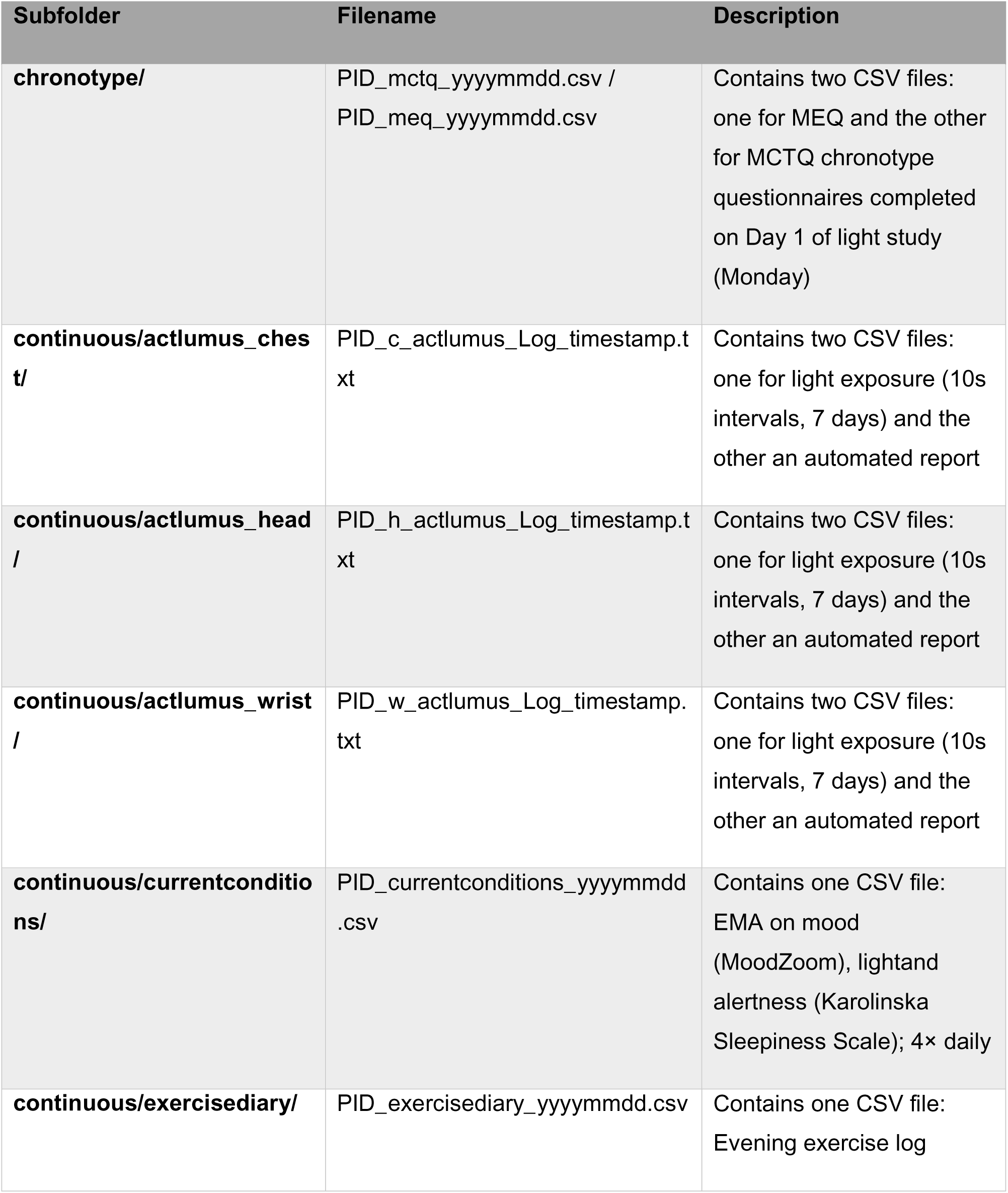
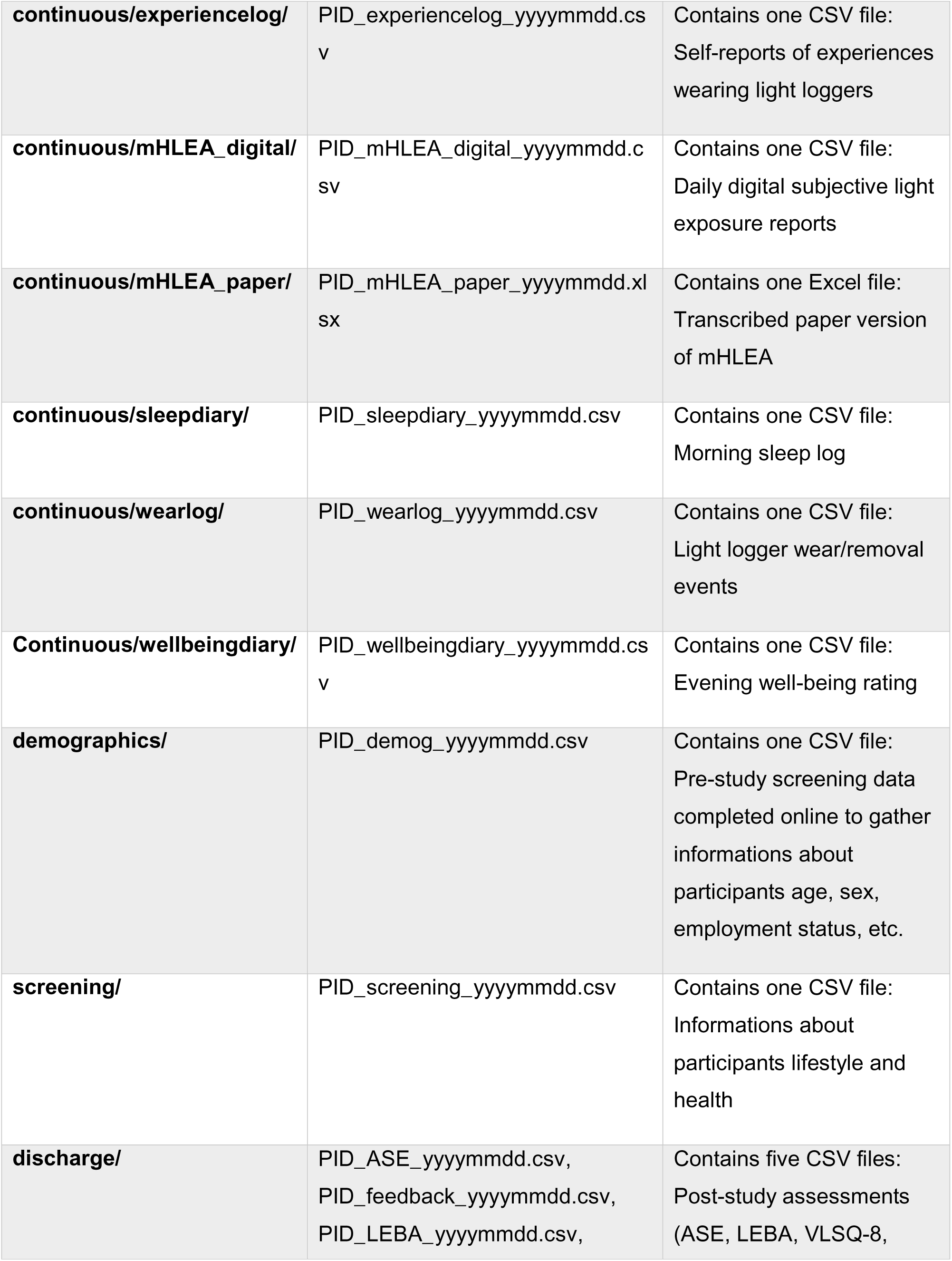
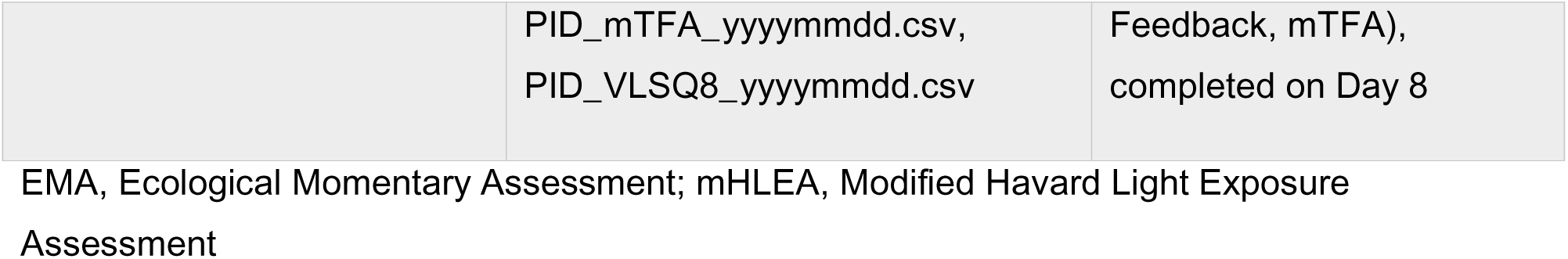
Description of individual-level folder.

## Data Availability

This dataset is publicly available on Zenodo under https://doi.org/10.5281/zenodo.15576732.

## Abbreviations

MCTQ: Munich Chronotype Questionnaire
MEQ: Morningness - Eveningness Questionnaire
mH-LEA: Modified Harvard Light Exposure Assessment
LEBA: Light Exposure Behaviour Assessment
VLSQ-8: Visual Light Sensitivity Questionnaire
ASE: Assessment of Sleep Environment
mTFA: Modified Theory Framework of Acceptability
EMA: Ecological Momentary Assessment
FAIR: Findable, Accessible, Interoperable and Reusable (data principles)
KNUST: Kwame Nkrumah University of Science and Technology
ID: Identification

## Statements

### Author contributions

Conceptualisation: GKA, JZ, MS, KOA

Data curation: GKA, JZ, KOA

Formal analysis: GKA, JZ, MS, KOA

Funding acquisition: MS, KOA

Investigation: GKA, MS, KOA

Methodology: GKA, JZ, MS, KOA

Project administration: GKA, KOA

Resources: GKA, IODJ, AKAA, JA, NAOM, AYAM, MS, KOA

Software: GKA, JZ, MS, KOA

Supervision: JZ, MS, KOA

Validation: GKA, JZ, MS, KOA

Visualisation: GKA, JZ, MS, KOA

Writing – original draft preparation: GKA, IODJ, AKAA, JA, NAOM, AYAM, KOA

Writing – review & editing: GKA, JZ, MS, IODJ, AKAA, JA, NAOM, AYAM, KOA

### Ethical approval

The multi-site study protocol was reviewed and approved by the Medical Ethics Committee of the Technical University of Munich (2024-118-S-SB). Locally, the study protocol was formally approved by the Committee on Human Research, Publication and Ethics at the School of Medicine and Dentistry, Kwame Nkrumah University of Science and Technology, Kumasi, Ghana (CHRPE/AP/644/24). Participants provided written informed consent, confirming their voluntary involvement in the research and their right to withdraw at any time, after the aims and approaches to be employed in the study had been thoroughly explained to them. All investigative procedures were performed in strict adherence to the Declaration of Helsinki.

## Acknowledgement

The authors would like to express gratitude to the participants who volunteered in this study, as well as Dr. Samson Darrah and Dr. Eldrick Adu Acquah for their unwavering technical assistance and support during the ethics application.

